# Lack of evidence for obesity paradox in patients with cardiovascular diseases: A UK BioBank cohort study

**DOI:** 10.1101/2025.01.07.25320160

**Authors:** Wenhao Yu, Bingbing Fan, Lin Yang, Shucheng Si, Wei Chen, Tao Zhang, Fuzhong Xue, Shengxu Li

**Affiliations:** Department of Biostatistics, School of Public Health, Cheeloo College of Medicine, Shandong University, Jinan, Shandong, China; Institute for Medical Dataology, Cheeloo College of Medicine, Shandong University, Jinan, Shandong, 250000, China; Department of Cancer Epidemiology and Prevention Research, Cancer Research & Analytics, Cancer Care Alberta, Alberta Health Services, Calgary, AB, Canada; Departments of Oncology and Community Health Sciences, Cumming School of Medicine, University of Calgary, Calgary, AB, Canada; Department of Epidemiology, Tulane University School of Public Health, New Orleans, Louisiana 70112; Department of Epidemiology and Biostatistics, School of Public Health, Tianjin Medical University, Tianjin, 300070, China; Children’s Minnesota Research Institute, Children’s Minnesota, Minneapolis, MN 55404

**Keywords:** obesity paradox, Mendelian randomization, cardiovascular disease, mortality

## Abstract

**Background:** Obesity paradox, a phenomenon that obesity seems to be associated with reduced risk of mortality in patients with established cardiovascular disease (CVD), has been controversial. We aimed to use Mendelian randomization to examine the causal relationship between obesity measures and CVD mortality in patients with known CVD in the UK BioBank study cohort.

**Methods:** A total of 58,278 participants with CVD were included. Polygenic risk scores (PRSs) for body mass index (BMI), body fat percentage (BF%), and waist to hip ration adjusted for BMI (WHRadjBMI) were used as instrumental variables. The following sensitivity analyses were performed: 1) using a representative variant rs1558902 in the fat mass and obesity associated gene as an instrumental variable, 2) by sex, and 3) by disease type.

**Results:** A total of 2203 patients died of CVD causes during a median follow-up period of 8.9 years. BMI in the overweight and class-I obesity range was associated with reduced mortality, with class-II or more severe obesity associated with increased mortality; however, there was a linear trend toward increased mortality with increasing BF% and WHRadjBMI. There was no clear indication that increased obesity-PRSs were associated with reduced risk of CVD mortality among individuals with known CVD. Sensitivity analyses using rs1558902 as an instrumental variable, by sex, and by disease type showed similar results.

**Conclusion:** Increased obesity does not show a protective effect in patients with CVD. Previously reported obesity paradox in observational studies may be a result of confounding or other biases, which needs further investigation.

## Introduction

Obesity is a major risk factor for cardiovascular disease (CVD)^1,2^. Large-scale meta-analyses have established the link between obesity and increased all-cause and CVD mortality^3,4^, and the causal relationship between obesity and CVD risk and associated mortality have been confirmed by recent Mendelian randomization studies^5–8^. As such, weight control is an important approach to mitigating burdens of CVD.

Despite the well-established relationship between obesity and CVD and mortality, there have been numerous reports showing that overweight or obesity is associated with better survival in patients with established CVD^9^. The phenomenon of apparent decreased risk associated with overweight/obesity, or rather high body mass index (BMI), has been termed “obesity paradox”. Notwithstanding large-scale meta-analyses showing the obesity paradox^10–15^, there have been contentious debates as to the very existence of the paradox or its clinical implications^16–18^. These debates center on causality and clinical implications of the paradox. One major argument against the existence of obesity paradox is that it is a result of complex confounding and bias, including collider bias and reverse causality^16^.

Mendelian randomization (MR) has become a go-to approach to examining causal relationships when direct evidence from randomized, controlled trials is lacking^19^. Large-scale genome-wide association studies have identified numerous genetic variants robustly associated with obesity measures, including BMI^20^, body fat percentage (BF%)^21^, and waist-to-hip ratio adjusted for BMI (WHRadjBMI)^22^. In a most recent study, Jenkins et al. adopted the MR approach and examined the causal relationship between BMI and mortality in patients with coronary heart disease, diabetes, or cancer, and did not find evidence to support obesity paradox^23^. However, the study focused solely on BMI, an obesity measure often criticized for its lack of ability to differentiate fat mass from muscle mass and to reflect fat distribution, and did not consider BF%, a direct adiposity measure. Further, the study did not consider disease types, such as heart failure, atrial fibrillation, and stroke. To address these limitations, the current Mendelian randomization study examined the genetic associations of BMI, BF%, and WHRadjBMI with mortality in patients with CVDs and the subtypes (when feasible) in the UK Biobank cohort.

## Methods

### Study population

This study was performed based on the UK Biobank cohort, a population-based cohort comprising around 500,000 people aged 40–69 years who were recruited from 2006 to 2010 across the UK. The cohort involves extensive phenotypic and genotypic details about its participants, including data from questionnaires, physical measures, sample assays, and genome-wide genotyping; it is linked to national medical records for longitudinal follow-up, including inpatient hospital episode records, primary care, cancer registries, and death registries^24^. After excluding individuals without genotypes, self-reported sex mismatched to genetic information, and >10 putative third-degree relatives in the kinship table, a total of 58,278 participants diagnosed with CVD diseases were included in the current study. Genetic quality control was performed centrally by the UK Biobank^25^. For detailed information about the cohort, please refer to its official website (https://www.ukbiobank.ac.uk/).

### Measurements

Anthropometric and laboratory data were collected by clinical technicians. Standing height, weight, waist, and hip circumference were measured. BMI was calculated as weight in kilograms divided by height in meters squared. BF% was measured by bioelectrical impedance using the Tanita BC418MA body composition analyzer. CVD in this research was defined by ICD-10 diagnostic codes from the health records of the UK Biobank. ICD-10 codes for all CVDs and type-specific CVDs were listed in **Supplementary Table S1**. Demographic and health behavioral variables, including age, sex, smoking, and drinking status, were self-reported. Smoking and drinking status was categorized into “Not to answer”, “Never”, “Previous”, and “Current”.

### Generation of polygenic risk scores (PRSs)

PRSs for BMI, BF%, and WHRadjBMI were generated for each participant from previously identified single nucleotide polymorphisms (SNPs) at the genome-wide significance levels (P < 5×10^−8^). The PRS for BMI (PRS-BMI) comprised 97 previously identified BMI variants (**Supplement Table S2**)^20^, the PRS for BF% (PRS-BF%) comprised 12 previously identified BF variants (**Supplement Table S3**)^21^, and the PRS for WHRadjBMI (PRS-WHRadjBMI) comprised 49 previously identified WHRadjBMI variants (**Supplement Table S4**)^22^. The PRSs for each individual participant were derived by summing the number of risk alleles that were each weighted by the allelic effect sizes (β-coefficients) published in the original GWAS meta-analysis^20–22^.

### Statistical analysis

Characteristics of study variables were compared using generalized linear models for continuous variables and X^2^ statistics for categorical variables. Five categories for weight status, underweight, normal weight, overweight, class-I obesity, and class-II obesity or above, were defined as BMI <18.5, 18.5-24.9, 25.0-29.9, 30.0-34.9, and ≥35.0 kg/m^2^, respectively. In addition to being used as a continuous variable, BF% and WHRadjBMI were also categorized into quintiles. We arbitrarily selected Q2 of BF% and Q1 of WHRadjBMI as the reference group. PRSs were also categorized into quintiles. Associations of obesity measures with quintiles of PRSs were evaluated for significance using a test for trend. Multiple logistic regression models were applied to examine the associations of PRSs with CVD mortality, with odds ratio (OR) and 95% confidence interval (CI) used to quantify the associations. Restricted cubic spline models were used to investigate the associations between obesity measures and CVD mortality by sex. To assess the robustness of associations, we additionally performed two-sample Mendelian randomization (TSMR) analyses^26^. We used the inverse-variance weighted^27^, MR-Egger^28^, weighted median^29^, and weighted mode-based method^30^ to estimate the causal effects. We also performed the following sensitivity analyses: 1) by sex; 2) by disease type (coronary heart disease, stroke, heart failure, atrial fibrillation, and peripheral artery disease); and 3) using a single SNP rs1558902 at the fat mass and obesity associated (*FTO*) gene as the instrument variable, to mitigate potential pleiotropic effects of the PRSs. The cardiometabolic consequences of common variants in the *FTO* gene closely mimic those of obesity^31^, and the SNPrs1558902 had the strongest associations with BMI in the study sample. All the statistical analyses were implemented with R version 4.1.0 and Plink 2 software.

## Results

A total of 58,278 participants with CVD were included in the current study (**Table 1**). The mean age of participants was 61.3 years, and 64.1% were males. Females had significantly higher hip circumference and BF%, but smaller waist circumference and lower proportions of smoking and drinking than males. As expected, there was a clear, strong increasing trend in BMI, BF%, and WHRadjBMI as the three PRSs increased from the bottom quintile to the top quintile in the total sample (**Figure 1**) and by sex (**Supplementary Figure S1**).

**Table 1.**
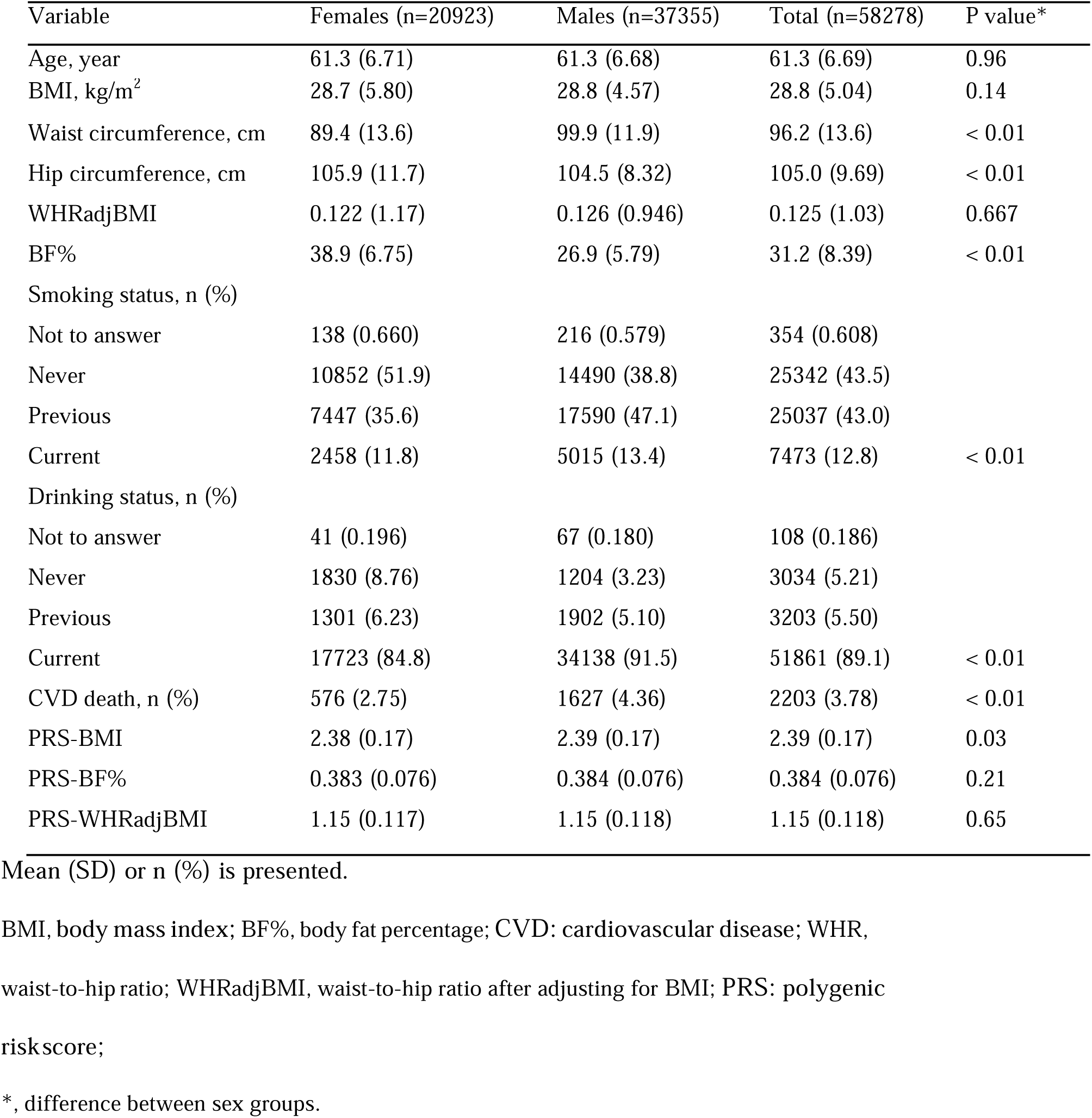
Characteristics of patients with cardiovascular and cerebrovascular diseases in the UK

| Variable | Females (n=20923) | Males (n=37355) | Total (n=58278) | P value* |
| --- | --- | --- | --- | --- |
| Age, year | 61.3 (6.71) | 61.3 (6.68) | 61.3 (6.69) | 0.96 |
| BMI, kg/m<sup>2</sup> | 28.7 (5.80) | 28.8 (4.57) | 28.8 (5.04) | 0.14 |
| Waist circumference, cm | 89.4 (13.6) | 99.9 (11.9) | 96.2 (13.6) | < 0.01 |
| Hip circumference, cm | 105.9 (11.7) | 104.5 (8.32) | 105.0 (9.69) | < 0.01 |
| WHRadjBMI | 0.122 (1.17) | 0.126 (0.946) | 0.125 (1.03) | 0.667 |
| BF% | 38.9 (6.75) | 26.9 (5.79) | 31.2 (8.39) | < 0.01 |
| Smoking status, n (%) |  |  |  |  |
| Not to answer | 138 (0.660) | 216 (0.579) | 354 (0.608) |  |
| Never | 10852 (51.9) | 14490 (38.8) | 25342 (43.5) |  |
| Previous | 7447 (35.6) | 17590 (47.1) | 25037 (43.0) |  |
| Current | 2458 (11.8) | 5015 (13.4) | 7473 (12.8) | < 0.01 |
| Drinking status, n (%) |  |  |  |  |
| Not to answer | 41 (0.196) | 67 (0.180) | 108 (0.186) |  |
| Never | 1830 (8.76) | 1204 (3.23) | 3034 (5.21) |  |
| Previous | 1301 (6.23) | 1902 (5.10) | 3203 (5.50) |  |
| Current | 17723 (84.8) | 34138 (91.5) | 51861 (89.1) | < 0.01 |
| CVD death, n (%) | 576 (2.75) | 1627 (4.36) | 2203 (3.78) | < 0.01 |
| PRS-BMI | 2.38 (0.17) | 2.39 (0.17) | 2.39 (0.17) | 0.03 |
| PRS-BF% | 0.383 (0.076) | 0.384 (0.076) | 0.384 (0.076) | 0.21 |
| PRS-WHRadjBMI | 1.15 (0.117) | 1.15 (0.118) | 1.15 (0.118) | 0.65 |

Mean (SD) or n (%) is presented.
BMI, body mass index; BF%, body fat percentage; CVD: cardiovascular disease; WHR, waist-to-hip ratio; WHRadjBMI, waist-to-hip ratio after adjusting for BMI; PRS: polygenic risk score;
\*, difference between sex groups.

**Figure 1.**
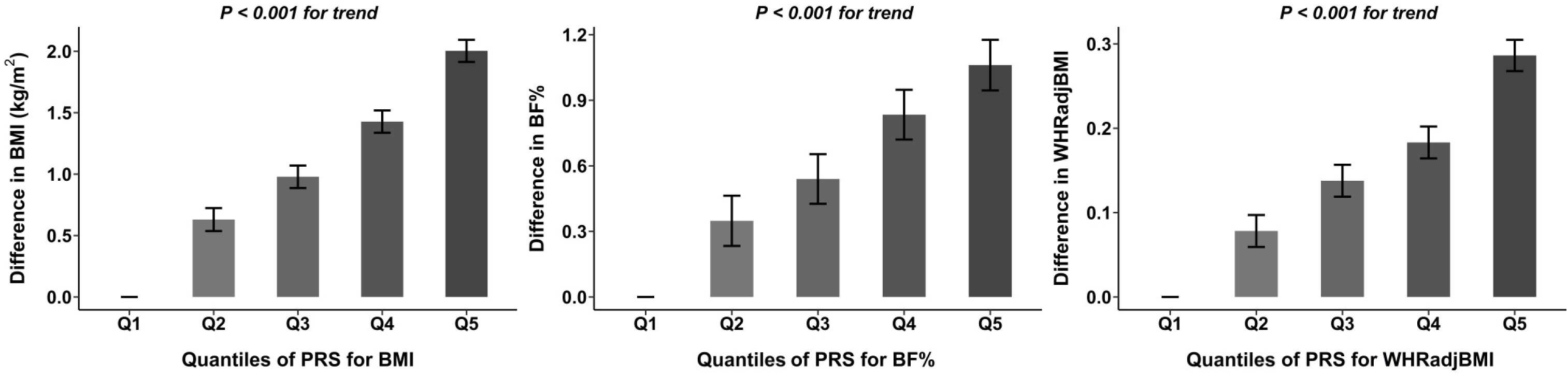
Associations of polygenic risk scores (PRSs) with BMI, BF%, and WHRadjBMI according to PRS quintiles BMI, body mass index; BF%, body fat percentage; WHRadjBMI, waist-to-hip ratio adjusted for BMI

A total of 2203 patients with CVD died of CVD causes during a median follow-up period of 8.9 (interquartile range 8.2-9.7) years. In a multiple logistic regression model with adjustment for age, sex, smoking and drinking status, compared with the normal weight group, the underweight group had the highest risk of CVD mortality (OR=2.62, 95% CI=1.52-4.24; P<0.01) and the overweight group had the lowest risk of CVD mortality (OR=0.86, 95% CI=0.76-0.97; P=0.01) (**Figure 1**). While the class-I obesity group did not show any reduction in CVD mortality (OR=0.97, 95% CI=0.85-1.10; P = 0.60), the class-II obesity or above group had significantly increased risk of CVD mortality (OR=1.49, 95% CI=1.28-1.73; P < 0.01) (**Figure 1**). Meanwhile, the associations of CVD mortality with BF% and WHRadjBMI appeared to be strengthening as the two obesity measures increased from the bottom quintile to the top quintile (**Figure 2**). The restricted cubic spline suggests a non-linear relationship between obesity measures and CVD mortality, and the associations were not the same for females and males (**Supplement Figure S2**). While U-shaped associations of the three obesity measures with CVD mortality were observed in males, an L-shaped association of BF%, and a J-shaped of WHRadjBMI, with CVD mortality in females were observed (**Supplementary Figure S2**).

**Figure 2.**
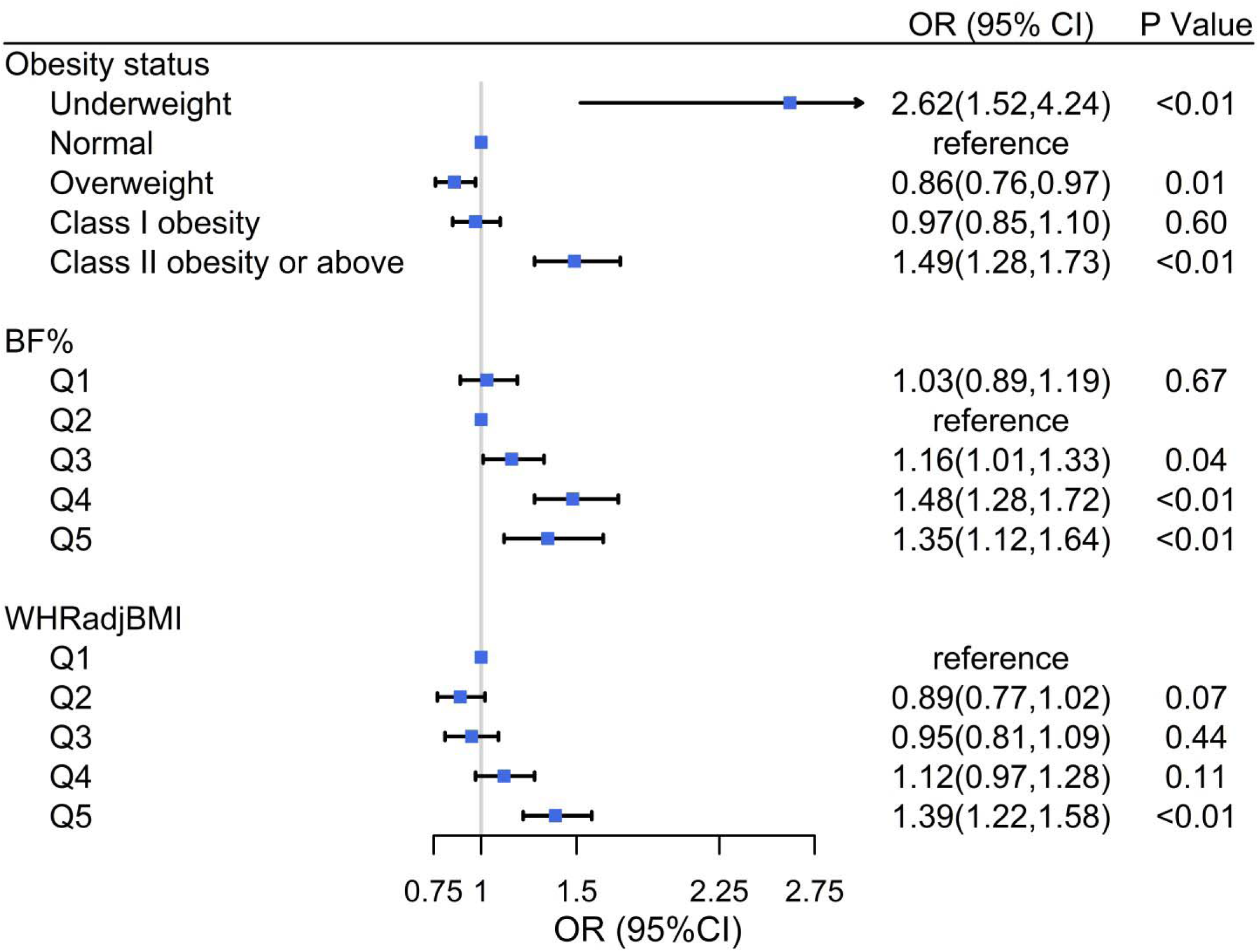
Associations of obesity measures with CVD mortality in patients with CVD. BF%, body fat percentage; BMI, body mass index; CI, confidence interval; OR, odds ratio; WHRadjBMI, waist-to-hip ratio adjusted for BMI Adjusted for age, sex, smoking and drinking status

After adjusting for age, sex, smoking and drinking status, there was no significantly lower risk of CVD mortality detected in higher quintile groups (ORs ranged from 0.88 to 1.10, P>0.05 for all), compared with the lowest quintile of PRSs (**Figure 3**). In two-sample MR analysis, there were no significant associations of the three PRSs with CVD mortality in patients with CVD (**Figure 4**, P>0.05 for all). Using rs1558902 in the *FTO* gene as the instrumental variable generated similar results (P>0.05).

**Figure 3.**
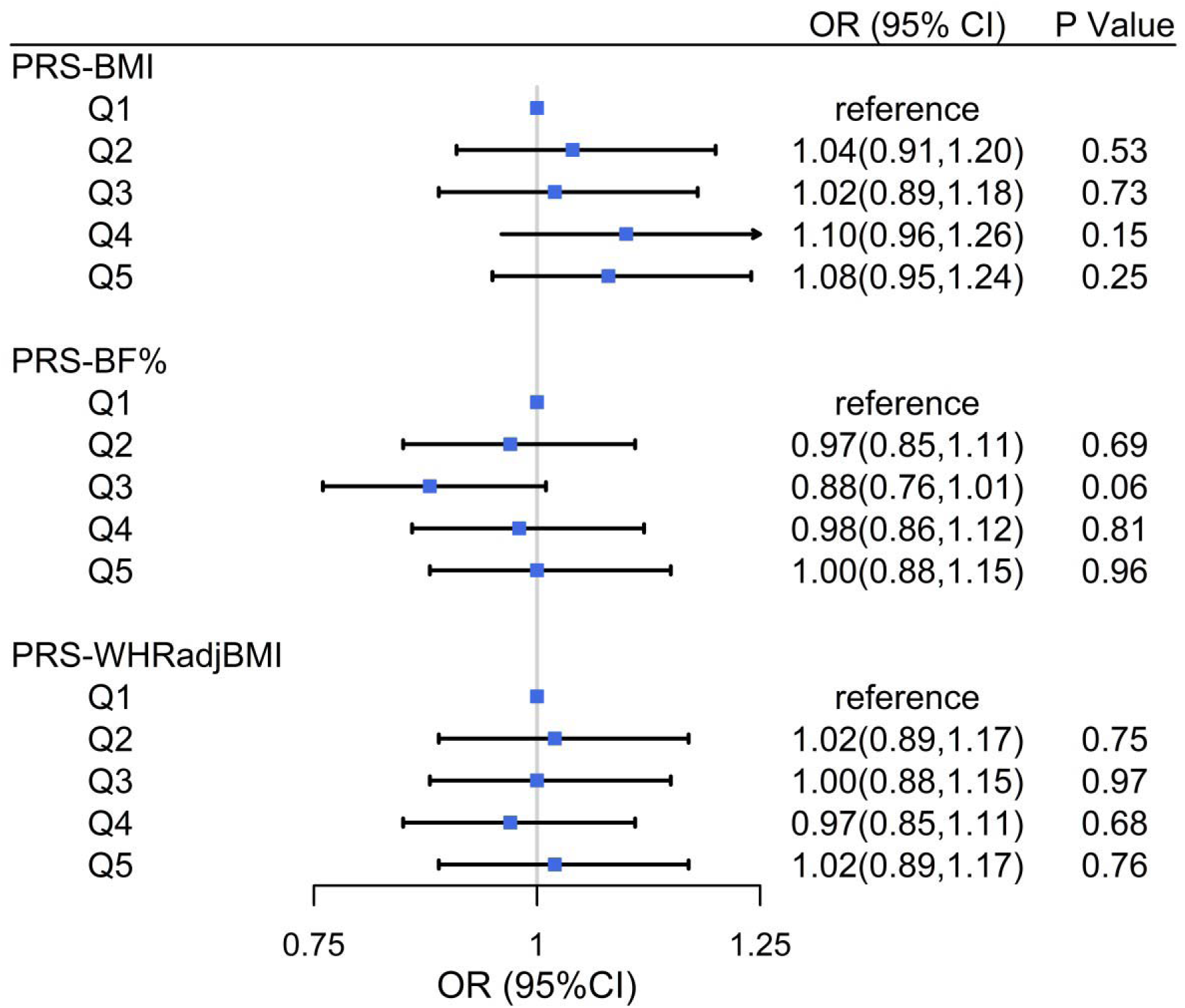
Associations of polygenetic risk scores (PRSs) with CVD mortality in patients with CVD BF%, body fat percentage; BMI, body mass index; CI, confidence interval; OR, odds ratio; WHRadjBMI, waist-to-hip ratio adjusted for BMI Adjusted for age, sex, smoking and drinking status

**Figure 4.**
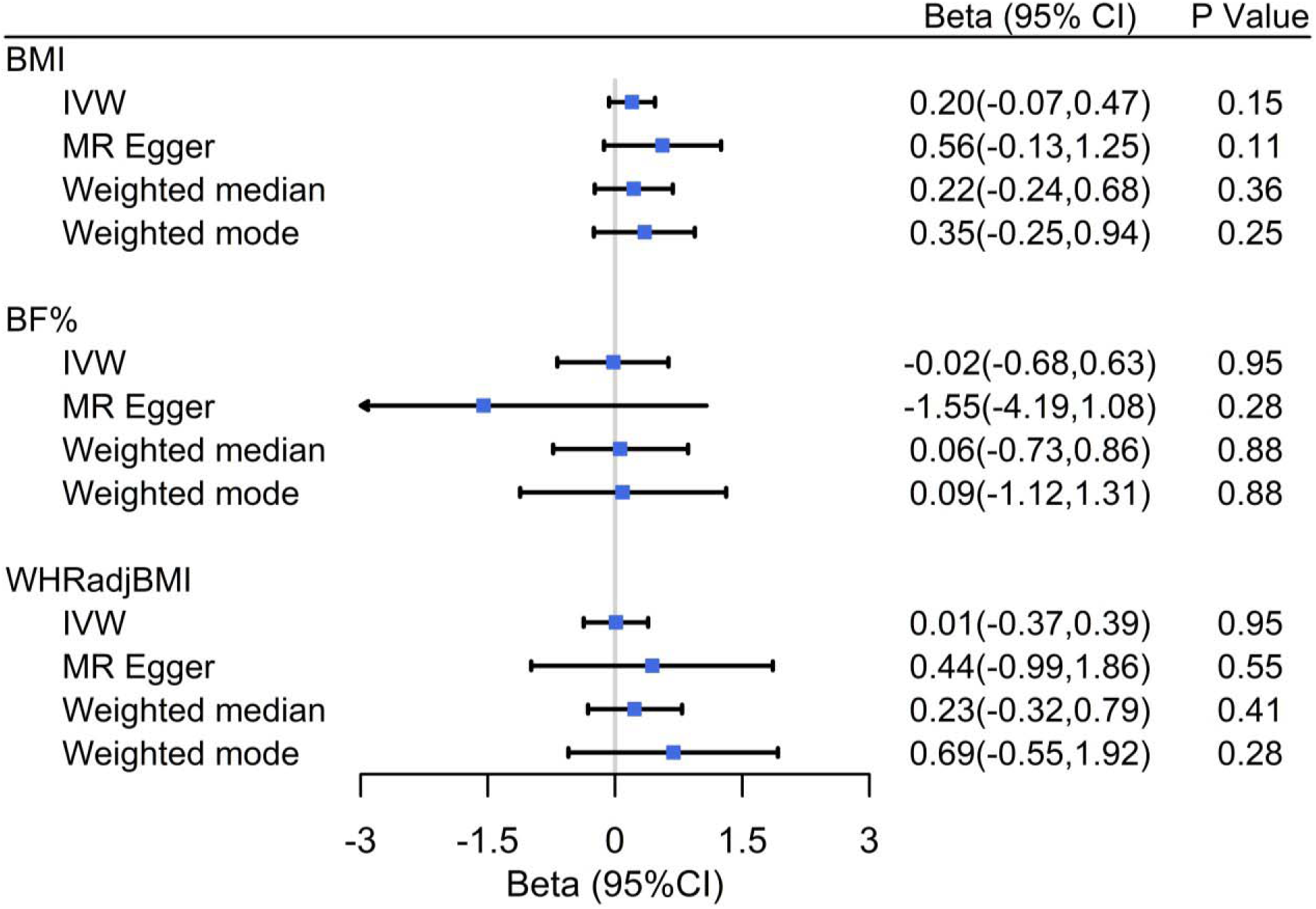
Two-sample Mendelian randomization analysis using polygenic risk scores for BMI, BF%, and WHRadjBMI as instrumental variables BF%, body fat percentage; BMI, body mass index; CI, confidence interval; IVW, inverse-variance weighted; WHRadjBMI, waist-to-hip ratio adjusted for BMI.

In sex-specific analyses, as shown in **Supplement Figures S3** and **S4**, except for the overweight [OR (95%CI): 0.80 (0.64,0.98), P=0.03] and class-I obesity group [OR (95%CI):0.69 (0.54,0.89), P<0.01] in females, obesity measures were not associated with CVD mortality. MR analyses did not show any associations of the 3 PRSs with CVD (**Supplement Figures S5** and **S6**). Results from two-sample MR analysis by sex (**Supplement Figure S7**) were consistent with results in the total sample in **Figure 4**. Results from disease-specific sensitivity analyses were overall consistent with results for the main analyses (**Supplementary Table S5**).

## Discussion

Findings of the current study do not support the existence of obesity paradox for CVD. While we did observe that CVD patients with overweight or obesity had reduced CVD mortality relative to that with normal weight, as previously observed for obesity paradox, the following results do not support the existence of obesity paradox: 1) BF%, a direct measure of adiposity, and WHRadjBMI, a measure of fat distribution, did not show evidence for obesity paradox; 2) increases in the three PRSs were not associated with a clear increasing trend in CVD mortality; and 3) lack of obesity paradox was corroborated by additional supplementary analyses by sex, by disease type, and by using the *FTO* variant as an instrument variable.

The fact that BF% and WHRadjBMI did not show any evidence of obesity paradox suggests that previous observations using BMI as a measure of obesity were likely biased or confounded. The evidence from MR also does not support the existence of obesity paradox. None of the three PRSs showed associations with CVD mortality in patients with CVD. Concerns on pleiotropic effects of the three PRSs are mitigated by using a single variant from the *FTO* gene as an instrument variable. Further, metabolic consequences of the variants from the *FTO* gene closely mimic those of obesity, including insulin resistance/type 2 diabetes, dyslipidemia, and hypertension^31^. Results using rs1558902 from the *FTO* gene did not support obesity paradox either. Despite sex discrepancies in body fatness and fat distribution, results from sex-specific analyses also did not support obesity paradox. We did not observe obesity paradox for individual outcomes including coronary heart disease, stroke, peripheral artery disease, and heart failure. In summary, all results from MR analyses did not support the existence of obesity paradox.

The current study significantly expanded the analyses conducted by Jenkins DA, et al^23^. We examined three obesity measures (BMI, BF%, and WHRadjBMI) whereas the Jenkins study only examined BMI. We also examined relationships between obesity measures and survival in different types of CVD. And finally, to mitigate concerns on pleiotropic effects of the PRSs, we also examined the associations of the *FTO* variant with CVD mortality, results of which were consistent with overall results.

Taken together the results from direct observations and from MR, our study does not support the existence of obesity paradox. Lack of obesity paradox is further supported by other secondary evidence. Patients with CVD or type 2 diabetes have significant reduced risk of mortality after bariatric surgery^32–34^. In addition, intentional weight loss is associated with improved heart function^15,35^. We hope that our study will help clarify the role of adiposity in survival of patients with CVD and dissipate the notion that increased body weight is a protective factor in patients with CVD. Nevertheless, we recognize the heterogeneity in etiology of CVD with and without obesity, which may have implications for tailored care for patients with CVD.

While the current study does not support obesity paradox, the question is what is behind previous observations of obesity paradox. Many hypotheses have been proposed, with explaining factors like confounding by age^36^, smoking^37,38^, physical fitness^39^, collider bias^40^, reverse causality^41^, differential care^41^, disease severity^38,42^, and even vascular regenerative capacity^43^. Although these hypotheses are plausible, none has yet to gain consistent literature support.

Another plausible explanation is that the relationship between obesity and CVD mortality has shifted such that the nadir for CVD and mortality is in the overweight or class I obesity range in the general population^44^, consistent with what is observed in a meta-analysis for heart failure^15^. A recent report suggests that obesity is a marker of less severe disease and does not have independent protective effect in patients with heart failure^42^, which needs to be confirmed in patients with other types of CVD. It has been suggested that obesity paradox in coronary artery disease may be a result of intact regenerative capacity in some, but not all, patients^43^. Taken together, the underlying causes for the observations of obesity paradox are yet to be proved with consistent and concrete evidence.

The current study has several strengths. The sample was from a population-based cohort and the sample size was large. The UK Biobank has rigorous quality assurance/control protocols. The main conclusion remained robust in a variety of sensitivity analyses, including sex-specific analysis, disease-specific MR analysis, and analysis using *FTO* variant rs1558902. As any other MR studies, the current study cannot eliminate the possibility of pleiotropy. Results from the single variant rs1558902, the metabolic consequences of which mimic those of obesity, however, still support our main conclusion. Another limitation is that CVD patients were prevalent patients, not incident patients, which might have introduced some biases associated with weight change due to the disease condition. However, obesity paradox is more easily observed in prevalent patients than in incident patients; lack of obesity paradox in prevalent patients argues for lack of obesity paradox.

In conclusion, the current study does not support the existence of obesity paradox. This conclusion helps clarify the role of adiposity in mortality risk in patients with CVD and support available guidelines on CVD prevention and intervention. A healthy diet and a physically active lifestyle should be promoted regardless of presence or absence of obesity in patients with CVD.

Underlying factors for the observations for obesity paradox remain unresolved.

## Data Availability

All data produced in the present study are available upon reasonable request to the authors.

https://www.ukbiobank.ac.uk/

## Acknowledgement

This work was partly supported by grants 82073572, 81973147, 82222064, and 82204124 from the National Natural Science Foundation of China.

