## Supplementary materials for "Lack of evidence for obesity paradox in patients with cardiovascular diseases: A UK BioBank cohort study"

Shandong, 250000, China

3 Department of Cancer Epidemiology and Prevention Research, Cancer Research & Analytics, Cancer Care Alberta, Alberta Health Services, Calgary, AB, Canada

4 Departments of Oncology and Community Health Sciences, Cumming School of Medicine,

University of Calgary, Calgary, AB, Canada

5 Department of Epidemiology, Tulane University School of Public Health, New Orleans, Louisiana 70112

6 Department of Epidemiology and Biostatistics, School of Public Health, Tianjin Medical University, Tianjin, 300070, China

7 Children’s Minnesota Research Institute, Children’s Minnesota, Minneapolis, MN 55404

### Address correspondence to:

Shengxu Li, MD, PhD, MPH

Children’s Minnesota Research Institute

Children’s Minnesota, MN 55404

Ema

Fuzhong Xue, MD, PhD

Department of Biostatistics, School of Public Health, Cheeloo College of Medicine, Shandong University, Jinan, Shandong, 250012, China

Institute for Medical Dataology, Cheeloo College of Medicine, Shandong University, Jinan, 250000, China

Ema

Tao Zhang, MD, PhD

Department of Biostatistics, School of Public Health, Cheeloo College of Medicine, Shandong University, Jinan, Shandong, 250012, China

Institute for Medical Dataology, Cheeloo College of Medicine, Shandong University, Jinan, 250000, China

Ema

**Supplement Table S1.** Definition of cardiovascular and cerebrovascular diseases in the UK Biobank.

| Disease | ICD10 in UK Biobank |
| --- | --- |
| CVD | I05, I06, I07, I08, I09, I10, I11, I12, I13, I15, I20, I21, I22, I23, I24, I25, I26, I27, I30, I33, I34, I35, I36, I37, I38, I42, I44, I46, I47, I48, I50, I60, I61, I62, I63, I64, I65, I66, I67, I68, I69, I70, I74, G45, G46, Q24 |
| Cardiovascular diseases | I05, I06, I07, I08, I09, I10, I11, I12, I13, I15, I20, I21, I22, I23, I24, I25, I26, I27, I30, I33, I34, I35, I36, I37, I38, I42, I44, I46, I47, I48, I50, I70, I74, Q24 |
| Stroke | I60, I61, I62, I63, I64, I65, I66, I67, I68, I69, G45, G46 |

**Supplement Table S2.** Individual 97 BMI single nucleotide polymorphism associations with BMI (in kg/m^2^)

| SNP | Chr:position | Alleles | EAF | Beta | SE | P |
| --- | --- | --- | --- | --- | --- | --- |
| rs657452 | 1:49362434 | A/G | 0.394 | 0.023 | 0.003 | 5.48E-13 |
| rs12286929 | 11:114527614 | G/A | 0.523 | 0.022 | 0.003 | 1.31E-12 |
| rs7903146 | 10:114748339 | C/T | 0.713 | 0.023 | 0.003 | 1.11E-11 |
| rs10132280 | 14:24998019 | C/A | 0.682 | 0.023 | 0.003 | 1.14E-11 |
| rs17094222 | 10:102385430 | C/T | 0.211 | 0.025 | 0.004 | 5.94E-11 |
| rs7599312 | 2:213121476 | G/A | 0.724 | 0.022 | 0.003 | 1.17E-10 |
| rs2365389 | 3:61211502 | C/T | 0.582 | 0.02 | 0.003 | 1.63E-10 |
| rs2820292 | 1:200050910 | C/A | 0.555 | 0.02 | 0.003 | 1.83E-10 |
| rs12885454 | 14:28806589 | C/A | 0.642 | 0.021 | 0.003 | 1.94E-10 |
| rs16851483 | 3:142758126 | T/G | 0.066 | 0.048 | 0.008 | 3.55E-10 |
| rs1167827 | 7:75001105 | G/A | 0.553 | 0.02 | 0.003 | 6.33E-10 |
| rs758747 | 16:3567359 | T/C | 0.265 | 0.023 | 0.004 | 7.47E-10 |
| rs1928295 | 9:119418304 | T/C | 0.548 | 0.019 | 0.003 | 7.91E-10 |
| rs9925964 | 16:31037396 | A/G | 0.62 | 0.019 | 0.003 | 8.11E-10 |
| rs11126666 | 2:26782315 | A/G | 0.283 | 0.021 | 0.003 | 1.33E-09 |
| rs2650492 | 16:28240912 | A/G | 0.303 | 0.021 | 0.004 | 1.92E-09 |
| rs6804842 | 3:25081441 | G/A | 0.575 | 0.019 | 0.003 | 2.48E-09 |
| rs4740619 | 9:15624326 | T/C | 0.542 | 0.018 | 0.003 | 4.56E-09 |
| rs13191362 | 6:162953340 | A/G | 0.879 | 0.028 | 0.005 | 7.34E-09 |
| rs3736485 | 15:49535902 | A/G | 0.454 | 0.018 | 0.003 | 7.41E-09 |
| rs17001654 | 4:77348592 | G/C | 0.153 | 0.031 | 0.005 | 7.76E-09 |
| rs11191560 | 10:104859028 | C/T | 0.089 | 0.031 | 0.005 | 8.45E-09 |
| rs1528435 | 2:181259207 | T/C | 0.631 | 0.018 | 0.003 | 1.20E-08 |
| rs1000940 | 17:5223976 | G/A | 0.32 | 0.019 | 0.003 | 1.28E-08 |
| rs2033529 | 6:40456631 | G/A | 0.293 | 0.019 | 0.003 | 1.39E-08 |
| rs11583200 | 1:50332407 | C/T | 0.396 | 0.018 | 0.003 | 1.48E-08 |
| rs9400239 | 6:109084356 | C/T | 0.688 | 0.019 | 0.003 | 1.61E-08 |
| rs10733682 | 9:128500735 | A/G | 0.478 | 0.017 | 0.003 | 1.83E-08 |
| rs11688816 | 2:62906552 | G/A | 0.525 | 0.017 | 0.003 | 1.89E-08 |
| rs11057405 | 12:121347850 | G/A | 0.901 | 0.031 | 0.006 | 2.02E-08 |
| rs11727676 | 4:145878514 | T/C | 0.91 | 0.036 | 0.006 | 2.55E-08 |
| rs3849570 | 3:81874802 | A/C | 0.359 | 0.019 | 0.003 | 2.60E-08 |
| rs6477694 | 9:110972163 | C/T | 0.365 | 0.017 | 0.003 | 2.67E-08 |
| rs7899106 | 10:87400884 | G/A | 0.052 | 0.04 | 0.007 | 2.96E-08 |
| rs2176598 | 11:43820854 | T/C | 0.251 | 0.02 | 0.004 | 2.97E-08 |
| rs2245368 | 7:76446079 | C/T | 0.18 | 0.032 | 0.006 | 3.19E-08 |
| rs17724992 | 19:18315825 | A/G | 0.746 | 0.019 | 0.004 | 3.42E-08 |
| rs7243357 | 18:55034299 | T/G | 0.812 | 0.022 | 0.004 | 3.86E-08 |
| rs2033732 | 8:85242264 | C/T | 0.747 | 0.019 | 0.004 | 4.89E-08 |
| rs9641123 | 7:93035668 | C/G | 0.43 | 0.029 | 0.005 | 2.08E-10 |
| rs7164727 | 15:70881044 | T/C | 0.671 | 0.019 | 0.003 | 3.92E-09 |
| rs492400 | 2:219057996 | C/T | 0.424 | 0.024 | 0.004 | 6.78E-09 |
| rs2080454 | 16:47620091 | C/A | 0.413 | 0.017 | 0.003 | 8.60E-09 |
| rs7239883 | 18:38401669 | G/A | 0.391 | 0.023 | 0.004 | 1.51E-08 |
| rs2836754 | 21:39213610 | C/T | 0.599 | 0.017 | 0.003 | 1.61E-08 |
| rs9914578 | 17:1951886 | G/C | 0.229 | 0.02 | 0.004 | 2.07E-08 |
| rs977747 | 1:47457264 | T/G | 0.403 | 0.017 | 0.003 | 2.18E-08 |
| rs9374842 | 6:120227364 | T/C | 0.744 | 0.023 | 0.004 | 2.67E-08 |
| rs4787491 | 16:29922838 | G/A | 0.51 | 0.022 | 0.004 | 2.70E-08 |
| rs1441264 | 13:78478920 | A/G | 0.613 | 0.017 | 0.003 | 2.96E-08 |
| rs17203016 | 2:207963763 | G/A | 0.195 | 0.021 | 0.004 | 3.41E-08 |
| rs16907751 | 8:81538012 | C/T | 0.913 | 0.047 | 0.009 | 3.89E-08 |
| rs13201877 | 6:137717234 | G/A | 0.14 | 0.024 | 0.004 | 4.29E-08 |
| rs9540493 | 13:65103705 | A/G | 0.452 | 0.021 | 0.004 | 4.97E-08 |
| rs1460676 | 2:164275935 | C/T | 0.179 | 0.021 | 0.004 | 4.98E-08 |
| rs6465468 | 7:95007450 | T/G | 0.306 | 0.025 | 0.005 | 4.98E-08 |
| rs6091540 | 20:50521269 | C/T | 0.721 | 0.03 | 0.004 | 2.15E-11 |
| rs7715256 | 5:153518086 | G/T | 0.422 | 0.017 | 0.003 | 8.85E-09 |
| rs2176040 | 2:226801046 | A/G | 0.365 | 0.024 | 0.004 | 9.99E-09 |
| rs1558902 | 16:52361075 | A/T | 0.415 | 0.082 | 0.003 | 7.51E-153 |
| rs6567160 | 18:55980115 | C/T | 0.236 | 0.056 | 0.004 | 3.93E-53 |
| rs13021737 | 2:622348 | G/A | 0.828 | 0.06 | 0.004 | 1.11E-50 |
| rs10938397 | 4:44877284 | G/A | 0.434 | 0.04 | 0.003 | 3.21E-38 |
| rs543874 | 1:176156103 | G/A | 0.193 | 0.048 | 0.004 | 2.62E-35 |
| rs2207139 | 6:50953449 | G/A | 0.177 | 0.045 | 0.004 | 4.13E-29 |
| rs11030104 | 11:27641093 | A/G | 0.792 | 0.041 | 0.004 | 5.56E-28 |
| rs3101336 | 1:72523773 | C/T | 0.613 | 0.033 | 0.003 | 2.66E-26 |
| rs7138803 | 12:48533735 | A/G | 0.384 | 0.032 | 0.003 | 8.15E-24 |
| rs10182181 | 2:25003800 | G/A | 0.462 | 0.031 | 0.003 | 8.78E-24 |
| rs3888190 | 16:28796987 | A/C | 0.403 | 0.031 | 0.003 | 3.14E-23 |
| rs1516725 | 3:187306698 | C/T | 0.872 | 0.045 | 0.005 | 1.89E-22 |
| rs12446632 | 16:19842890 | G/A | 0.865 | 0.04 | 0.005 | 1.48E-18 |
| rs2287019 | 19:50894012 | C/T | 0.804 | 0.036 | 0.004 | 4.59E-18 |
| rs16951275 | 15:65864222 | T/C | 0.784 | 0.031 | 0.004 | 1.91E-17 |
| rs3817334 | 11:47607569 | T/C | 0.407 | 0.026 | 0.003 | 5.15E-17 |
| rs2112347 | 5:75050998 | T/G | 0.629 | 0.026 | 0.003 | 6.19E-17 |
| rs12566985 | 1:74774781 | G/A | 0.446 | 0.024 | 0.003 | 3.28E-15 |
| rs3810291 | 19:52260843 | A/G | 0.666 | 0.028 | 0.004 | 4.81E-15 |
| rs7141420 | 14:78969207 | T/C | 0.527 | 0.024 | 0.003 | 1.23E-14 |
| rs13078960 | 3:85890280 | G/T | 0.196 | 0.03 | 0.004 | 1.74E-14 |
| rs10968576 | 9:28404339 | G/A | 0.32 | 0.025 | 0.003 | 6.61E-14 |
| rs17024393 | 1:109956211 | C/T | 0.04 | 0.066 | 0.009 | 7.03E-14 |
| rs12429545 | 13:53000207O | A/G | 0.133 | 0.033 | 0.005 | 1.09E-12 |
| rs13107325 | 4:103407732 | T/C | 0.072 | 0.048 | 0.007 | 1.83E-12 |
| rs11165643 | 1:96696685 | T/C | 0.583 | 0.022 | 0.003 | 2.07E-12 |
| rs17405819 | 8:76969139 | T/C | 0.7 | 0.022 | 0.003 | 2.07E-11 |
| rs1016287 | 2:59159129 | T/C | 0.287 | 0.023 | 0.003 | 2.25E-11 |
| rs4256980 | 11:8630515 | G/C | 0.646 | 0.021 | 0.003 | 2.90E-11 |
| rs12401738 | 1:78219349 | A/G | 0.352 | 0.021 | 0.003 | 1.15E-10 |
| rs205262 | 6:34671142 | G/A | 0.273 | 0.022 | 0.004 | 1.75E-10 |
| rs12016871 | 13:26915782 | T/C | 0.203 | 0.03 | 0.005 | 2.29E-10 |
| rs12940622 | 17:76230166 | G/A | 0.575 | 0.018 | 0.003 | 2.49E-09 |
| rs11847697 | 14:29584863 | T/C | 0.042 | 0.049 | 0.008 | 3.99E-09 |
| rs2075650 | 19:50087459 | A/G | 0.848 | 0.026 | 0.005 | 1.25E-08 |
| rs2121279 | 2:142759755 | T/C | 0.152 | 0.025 | 0.004 | 2.31E-08 |
| rs29941 | 19:39001372 | G/A | 0.669 | 0.018 | 0.003 | 2.41E-08 |
| rs1808579 | 18:19358886 | C/T | 0.534 | 0.017 | 0.003 | 4.17E-08 |

SNP, single nucleotide polymorphism; Chr, chromosome; EAF, effect allele frequency.

**Supplement Table S3.** Individual 12 body fat single nucleotide polymorphism associations with BF (%)

| SNP | Chr | Locus | EA | EAF | Beta | SE | P |
| --- | --- | --- | --- | --- | --- | --- | --- |
| rs1558902 | 16 | 52361075 | A | 40% | 0.051 | 0.005 | 3.80E-27 |
| rs2943652 | 2 | 226816690 | C | 36% | 0.034 | 0.005 | 1.50E-12 |
| rs6567160 | 18 | 55980115 | C | 25% | 0.034 | 0.005 | 1.30E-10 |
| rs6755502 | 2 | 625721 | C | 83% | 0.039 | 0.006 | 1.40E-10 |
| rs6738627 | 2 | 165252696 | A | 37% | 0.03 | 0.005 | 5.70E-09 |
| rs693839 | 13 | 79856289 | C | 32% | 0.028 | 0.005 | 6.60E-09 |
| rs6857 | 19 | 50084094 | C | 83% | 0.048 | 0.008 | 6.80E-09 |
| rs4788099 | 16 | 28763228 | G | 38% | 0.027 | 0.005 | 1.20E-08 |
| rs9906944 | 17 | 44446419 | C | 67% | 0.033 | 0.006 | 2.90E-08 |
| rs543874 | 1 | 176156103 | G | 19% | 0.032 | 0.006 | 4.50E-08 |
| rs3761445 | 22 | 36925357 | G | 41% | 0.024 | 0.005 | 1.70E-07 |
| rs757318 | 19 | 18681308 | C | 50% | 0.024 | 0.005 | 2.10E-07 |

SNP, single nucleotide polymorphism; Chr, chromosome; EA, effect allele; EAF, effect allele frequency.

**Supplement Table S4.** Individual 49 single nucleotide polymorphism associations with WHRadjBMI

| SNP | Chr | Position | EA | EAF | Beta | P |
| --- | --- | --- | --- | --- | --- | --- |
| rs905938 | 1 | DCST2 | T | 0.74 | 0.025 | 7.30E-01 |
| rs10919388 | 1 | GORAB | C | 0.72 | 0.024 | 3.20E-09 |
| rs1385167 | 2 | MEIS1 | G | 0.15 | 0.029 | 1.90E-09 |
| rs1569135 | 2 | CALCRL | A | 0.53 | 0.021 | 5.60E-01 |
| rs10804591 | 3 | PLXND1 | A | 0.79 | 0.025 | 6.60E-09 |
| rs17451107 | 3 | LEKR1 | T | 0.61 | 0.026 | 1.10E-12 |
| rs3805389 | 4 | NMU | A | 0.28 | 0.012 | 1.50E-03 |
| rs9991328 | 4 | FAM13A | T | 0.49 | 0.019 | 4.50E-08 |
| rs303084 | 4 | SPATA5-FGF2 | A | 0.8 | 0.023 | 3.90E-08 |
| rs9687846 | 5 | MAP3K1 | A | 0.19 | 0.024 | 7.10E-08 |
| rs6556301 | 5 | FGFR4 | T | 0.36 | 0.022 | 2.60E-08 |
| rs7759742 | 6 | BTNL2 | A | 0.51 | 0.023 | 4.40E-11 |
| rs1776897 | 6 | HMGA1 | G | 0.08 | 0.03 | 1.10E-05 |
| rs7801581 | 7 | HOXA11 | T | 0.24 | 0.027 | 3.70E-01 |
| rs7830933 | 8 | NKX2-6 | A | 0.77 | 0.022 | 7.40E-08 |
| rs12679556 | 8 | MSC | G | 0.25 | 0.027 | 2.10E-11 |
| rs10991437 | 9 | ABCA1 | A | 0.11 | 0.031 | 1.00E-08 |
| rs7917772 | 10 | SFXN2 | A | 0.62 | 0.014 | 5.60E-05 |
| rs11231693 | 11 | MACROD1-VEGFB | A | 0.06 | 0.041 | 4.50E-08 |
| rs4765219 | 12 | CCDC92 | C | 0.67 | 0.028 | 1.60E-15 |
| rs8042543 | 15 | KLF13 | C | 0.78 | 0.026 | 1.20E-09 |
| rs8030605 | 15 | RFX7 | A | 0.14 | 0.03 | 8.80E-09 |
| rs1440372 | 15 | SMAD6 | C | 0.71 | 0.024 | 1.10E-01 |
| rs2925979 | 16 | CMIP | T | 0.31 | 0.018 | 1.20E-06 |
| rs4646404 | 17 | PEMT | G | 0.67 | 0.027 | 1.40E-11 |
| rs8066985 | 17 | KCNJ2 | A | 0.5 | 0.018 | 1.40E-07 |
| rs12454712 | 18 | BCL2 | T | 0.61 | 0.016 | 1.00E-04 |
| rs12608504 | 19 | JUND | A | 0.36 | 0.022 | 8.80E-01 |
| rs4081724 | 19 | CEBPA | G | 0.85 | 0.035 | 7.40E-12 |
| rs979012 | 20 | BMP2 | T | 0.34 | 0.027 | 3.30E-14 |
| rs224333 | 20 | GDF5 | G | 0.62 | 0.02 | 2.60E-08 |
| rs6090583 | 20 | EYA2 | A | 0.48 | 0.022 | 6.20E-11 |
| rs1534696 | 7 | SNX10 | C | 0.43 | 0.011 | 1.30E-03 |
| rs2645294 | 1 | TBX15-WARS2 | T | 0.58 | 0.031 | 1.70E-19 |
| rs714515 | 1 | DNM3-PIGC | G | 0.43 | 0.027 | 4.40E-15 |
| rs2820443 | 1 | LYPLAL1 | T | 0.72 | 0.035 | 5.30E-21 |
| rs10195252 | 2 | GRB14-COBLL1 | T | 0.59 | 0.027 | 5.90E-15 |
| rs17819328 | 3 | PPARG | G | 0.43 | 0.021 | 2.40E-09 |
| rs2276824 | 3 | PBRM1 | C | 0.43 | 0.024 | 3.20E-11 |
| rs2371767 | 3 | ADAMTS9 | G | 0.72 | 0.036 | 1.60E-02 |
| rs1045241 | 5 | TNFAIP8-HSD17B4 | C | 0.71 | 0.019 | 4.40E-07 |
| rs7705502 | 5 | CPEB4 | A | 0.33 | 0.027 | 4.70E-14 |
| rs1294410 | 6 | LY86 | C | 0.63 | 0.031 | 2.00E-18 |
| rs1358980 | 6 | VEGFA | T | 0.47 | 0.039 | 3.10E-27 |
| rs1936805 | 6 | RSPO3 | T | 0.51 | 0.043 | 3.60E-35 |
| rs10245353 | 7 | NFE2L3 | A | 0.2 | 0.035 | 8.40E-16 |
| rs10842707 | 12 | ITPR2-SSPN | T | 0.23 | 0.032 | 4.40E-16 |
| rs1443512 | 12 | HOXC13 | A | 0.24 | 0.028 | 6.90E-13 |
| rs2294239 | 22 | ZNRF3 | A | 0.59 | 0.025 | 7.20E-13 |

SNP, single nucleotide polymorphism; Chr, chromosome; EA, effect allele; EAF, effect allele frequency.

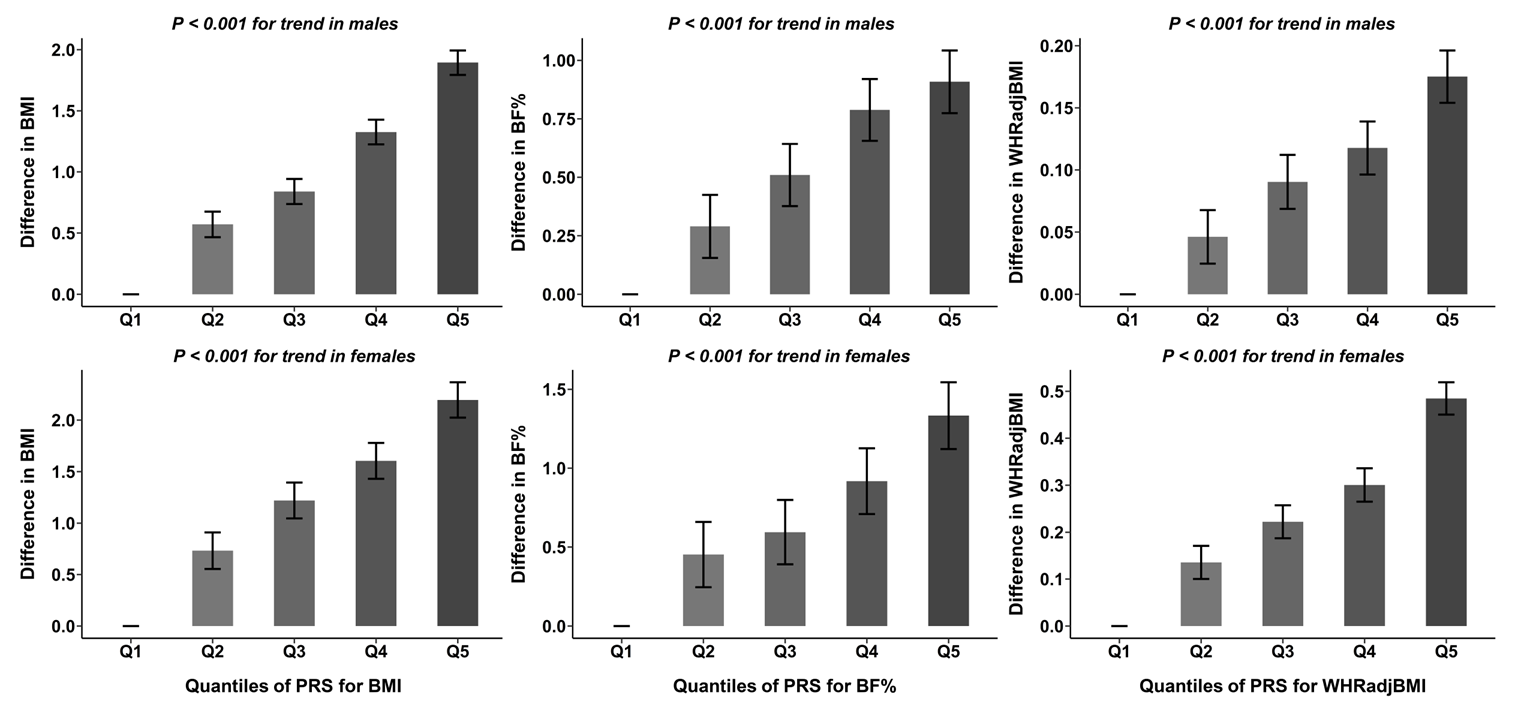

**Supplement Figure S1.** BMI, BF%, and WHRadjBMI by quintile of polygenic risk scores (PRS)

BF%, body fat percentage; BMI, body mass index; WHRadjBMI, waist-to-hip ratio adjusted for BMI.

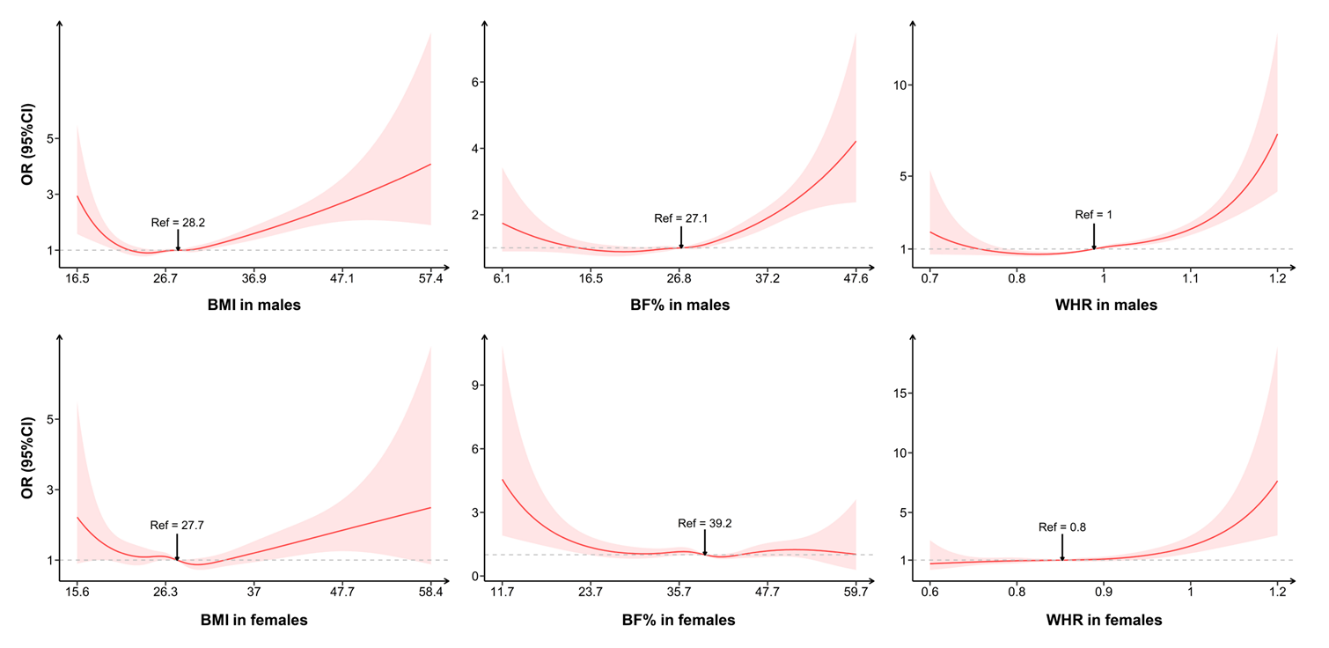

**Supplement Figure S2.** Associations of BMI, BF%, and WHR with CVD death based on restricted cubic spline by sex

BF%, body fat percentage; BMI, body mass index; WHRadjBMI, waist-to-hip ratio adjusted for BMI. Adjusted for age, smoking, and drinking status

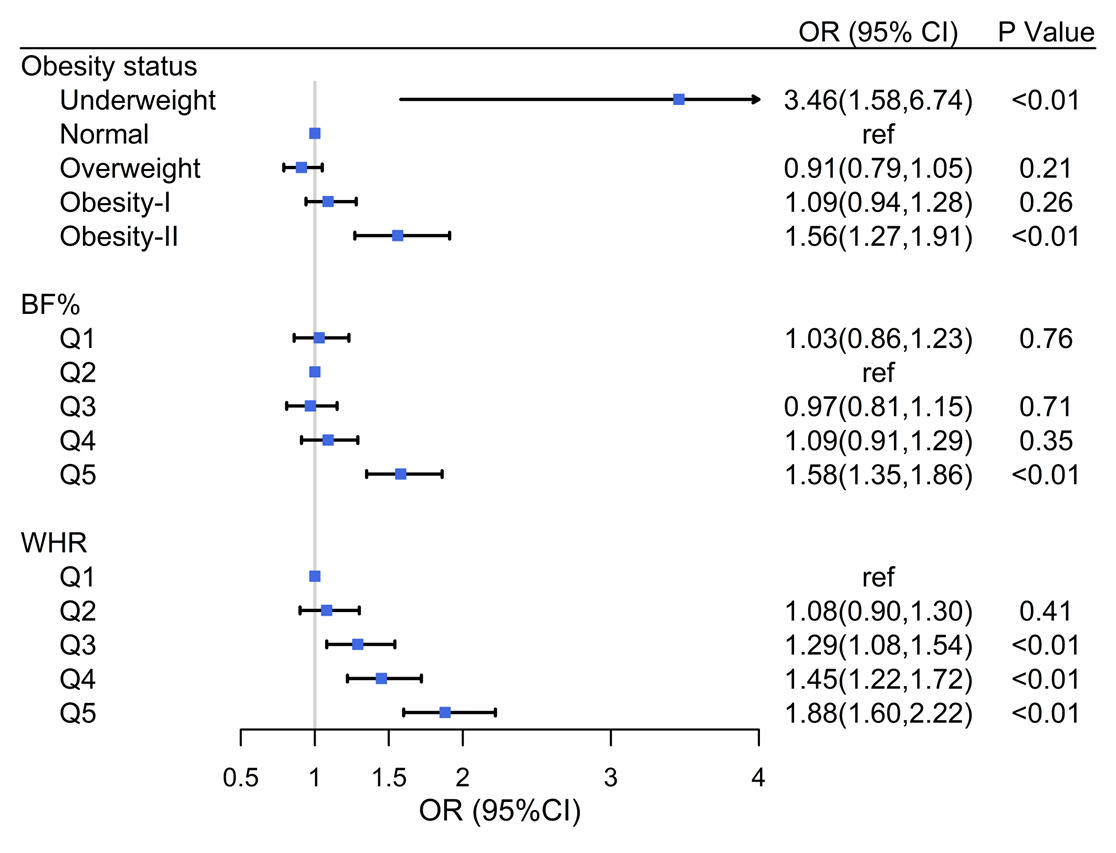

**Supplement Figure S3.** Associations of obesity status, BF%, WHRadjBMI with CVD death in **males** with CVD

BF%, body fat percentage; CI, 95% confidence interval; OR, odds ratio; WHRadjBMI, waist-to-hip ratio

Adjusted for age, smoking and drinking status.

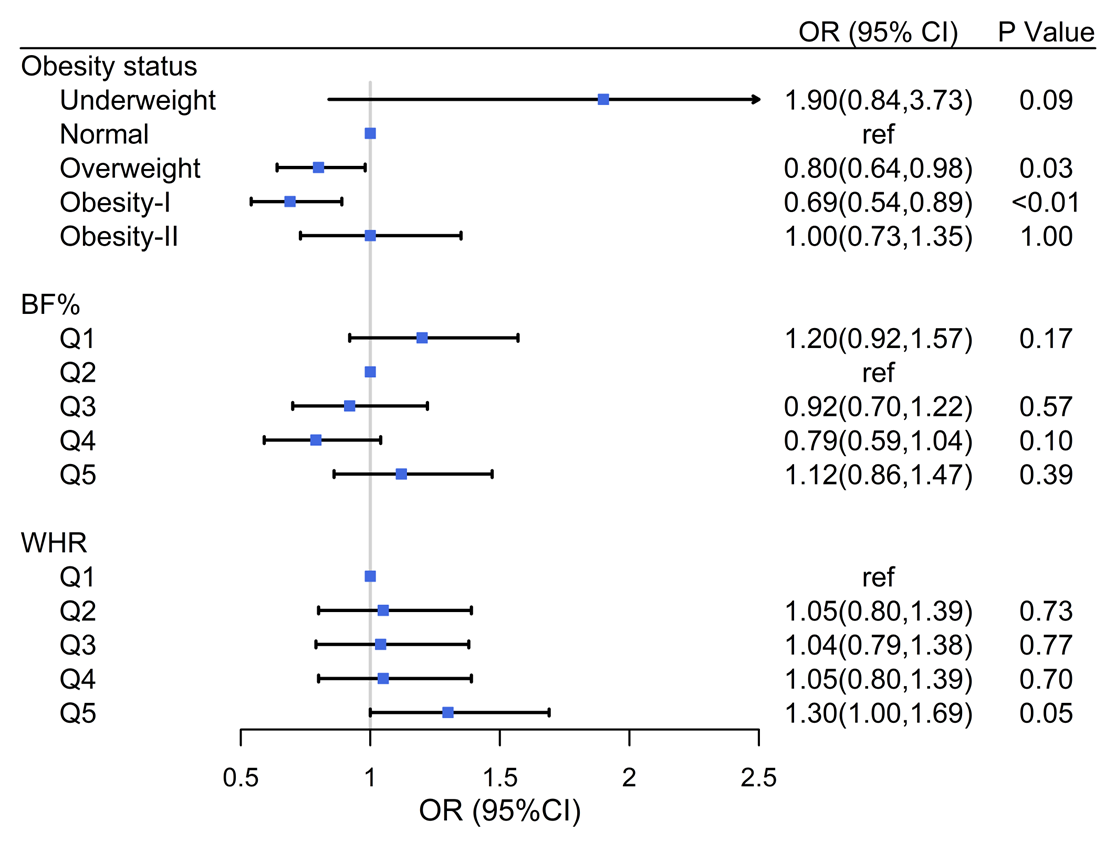

**Supplement Figure S4.** Associations of obesity status, BF%, WHRadjBMI with CVD death in **females** with CVD

BF%, body fat percentage; CI, 95% confidence interval; OR, odds ratio; WHRadjBMI, waist-to-hip ratio

Adjusted for age, smoking and drinking status.

.

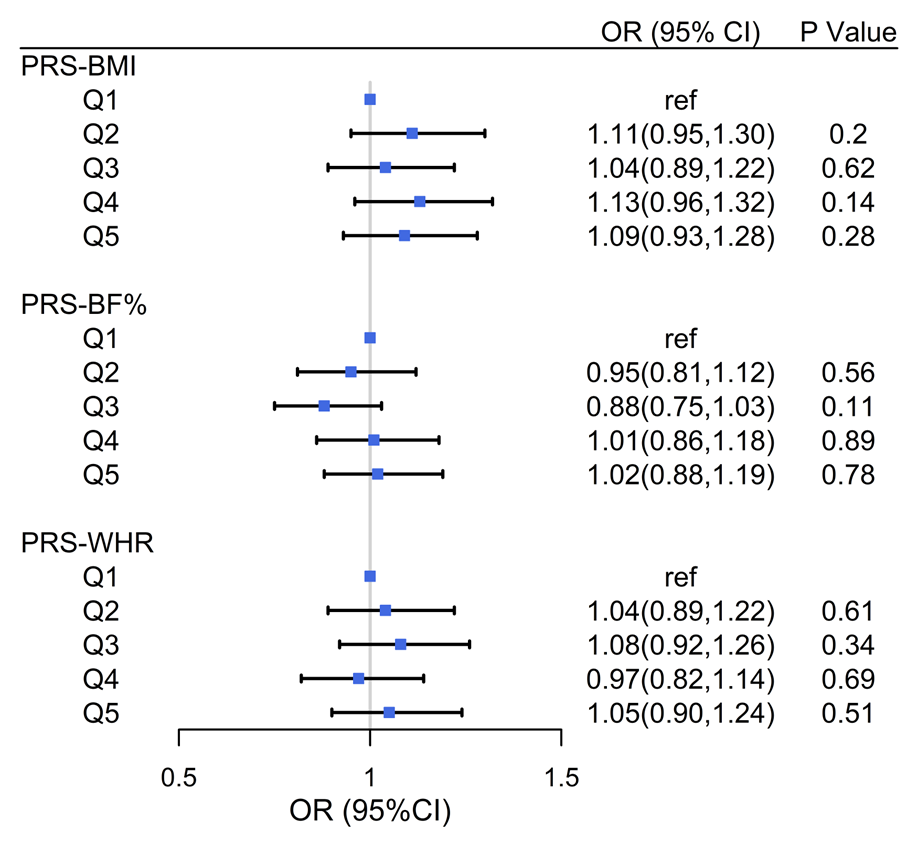

**Supplement Figure S5.** Associations of polygenic risk scores (PRSs) with CVD death in **males** with CVD

BF%, body fat percentage; CI, 95% confidence interval; OR, odds ratio; WHRadjBMI, waist-to-hip ratio

Adjusted for age, smoking and drinking status.

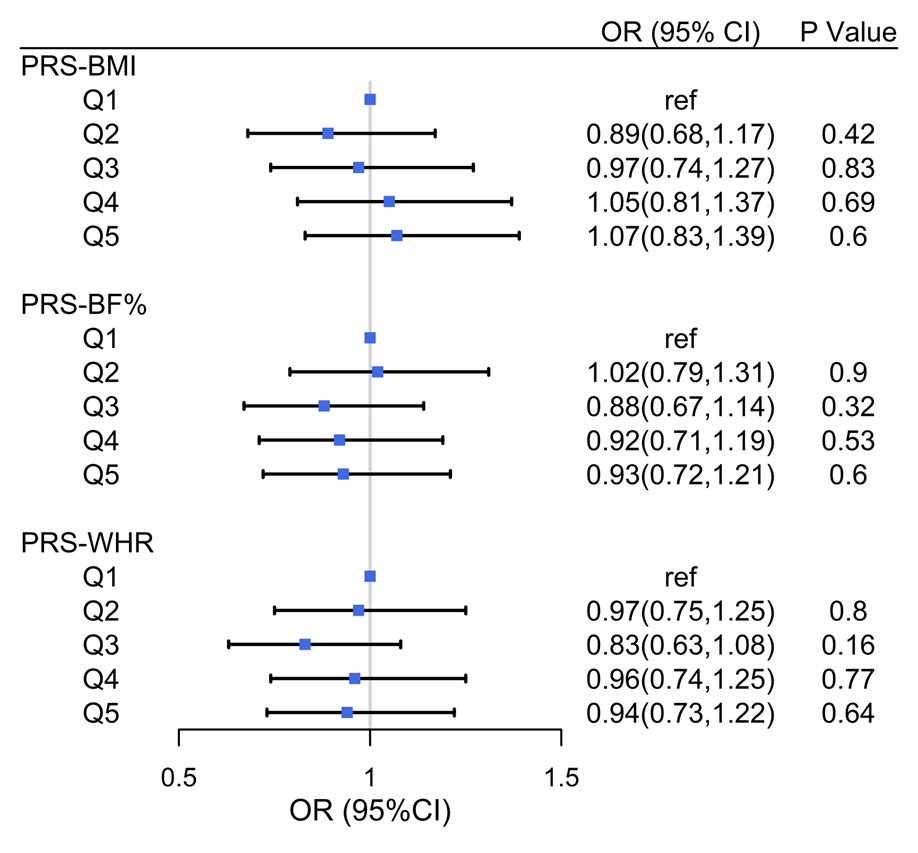

**Supplement Figure S6.** Associations of polygenic risk scores (PRSs) with CVD death in **females** with CVD

BF%, body fat percentage; CI, 95% confidence interval; OR, odds ratio; WHRadjBMI, waist-to-hip ratio

Adjusted for age, smoking and drinking status.

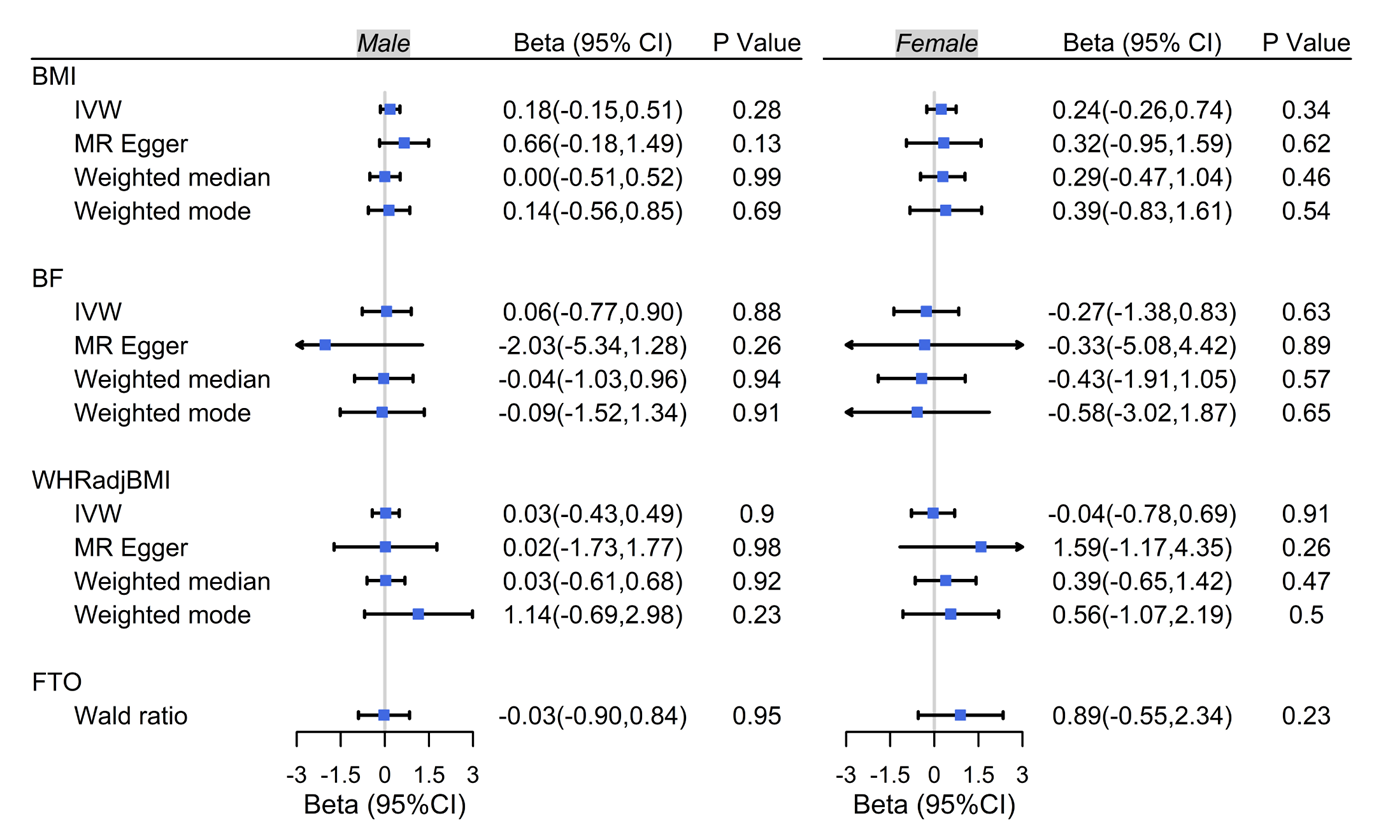

**Supplement Figure S7.** Two-sample Mendelian randomization analysis by sex

BF%, body fat percentage; BMI, body mass index; CI, 95% confidence interval; IVW, inverse-variance weighted; OR, odds ratio; WHRadjBMI, waist-to-hip ratio

Adjusted for age, smoking and drinking status.

**Supplement Table S5.** Associations of BMI, BF%, and WHRadjBMI, and their respective PRSs with specific CVD mortality

|  | CHD |  |  | Heart Failure |  |  | Stroke |  |  | Atrial fibrillation |  |  | PAD |  |
| --- | --- | --- | --- | --- | --- | --- | --- | --- | --- | --- | --- | --- | --- | --- |
|  | OR (95%CI) | P |  | OR (95%CI) | P |  | OR (95%CI) | P |  | OR (95%CI) | P value |  | OR (95%CI) | P |
| Obesity status |  |  |  |  |  |  |  |  |  |  |  |  |  |  |
| Underweight | 1.82 (0.55, 4.47) | 0.253 |  | 2.71 (0.81, 8.05) | 0.082 |  | 2.02 (0.85, 4.27) | 0.085 |  | 4.61 (1.06, 13.9) | 0.016 |  | 1.32E-6 (0, 3.94E+18) | 0.982 |
| Normal | reference |  |  | reference |  |  | reference |  |  | reference |  |  | reference |  |
| Overweight | 0.94 (0.79, 1.12) | 0.463 |  | 0.98 (0.72, 1.37) | 0.926 |  | 0.80 (0.66, 0.98) | 0.032 |  | 0.90 (0.66, 1.24) | 0.492 |  | 0.71 (0.37, 1.41) | 0.318 |
| Obesity-I | 1.09 (0.91, 1.32) | 0.355 |  | 0.99 (0.71, 1.39) | 0.954 |  | 0.82 (0.65, 1.03) | 0.094 |  | 1.25 (0.90, 1.75) | 0.189 |  | 0.96 (0.48, 1.98) | 0.922 |
| Obesity-II | 1.49 (1.18, 1.89) | 0.001 |  | 1.16 (0.79, 1.69) | 0.458 |  | 0.94 (0.66, 1.31) | 0.720 |  | 2.21 (1.50, 3.23) | < 0.001 |  | 0.60 (0.16, 1.74) | 0.380 |
| BF% |  |  |  |  |  |  |  |  |  |  |  |  |  |  |
| Q1 | 1.08 (0.88, 1.32) | 0.473 |  | 0.93 (0.66, 1.32) | 0.690 |  | 1.04 (0.80, 1.36) | 0.765 |  | 1.20 (0.81, 1.79) | 0.358 |  | 0.69 (0.28, 1.66) | 0.418 |
| Q2 | reference |  |  | reference |  |  | reference |  |  | reference |  |  | reference |  |
| Q3 | 1.29 (1.06, 1.57) | 0.010 |  | 0.82 (0.58, 1.16) | 0.268 |  | 1.06 (0.82, 1.38) | 0.650 |  | 1.33 (0.91, 1.97) | 0.142 |  | 0.73 (0.30, 1.71) | 0.469 |
| Q4 | 1.63 (1.34, 1.99) | < 0.001 |  | 1.18 (0.83, 1.67) | 0.364 |  | 1.05 (0.79, 1.39) | 0.736 |  | 2.54 (1.75, 3.700) | < 0.001 |  | 1.57 (0.73, 3.43) | 0.246 |
| Q5 | 1.64 (1.24, 2.16) | < 0.001 |  | 0.86 (0.54, 1.35) | 0.514 |  | 0.84 (0.60, 1.16) | 0.292 |  | 1.82 (1.09, 3.04) | 0.022 |  | 1.69 (0.63, 4.45) | 0.290 |
| WHR |  |  |  |  |  |  |  |  |  |  |  |  |  |  |
| Q1 | reference |  |  | reference |  |  | reference |  |  | reference |  |  | reference |  |
| Q2 | 1.07 (0.84, 1.37) | 0.589 |  | 1.56 (1.08, 2.27) | 0.017 |  | 0.79 (0.60, 1.02) | 0.075 |  | 0.96 (0.60, 1.53) | 0.850 |  | 1.39 (0.55, 3.66) | 0.492 |
| Q3 | 1.29 (1.01, 1.66) | 0.043 |  | 1.36 (0.91, 2.02) | 0.132 |  | 0.78 (0.58, 1.04) | 0.094 |  | 1.39 (0.88, 2.21) | 0.165 |  | 1.04 (0.38, 2.96) | 0.938 |
| Q4 | 1.53 (1.19, 1.97) | 0.001 |  | 1.44 (0.96, 2.15) | 0.076 |  | 0.88 (0.65, 1.19) | 0.402 |  | 1.52 (0.96, 2.45) | 0.078 |  | 1.44 (0.56, 3.93) | 0.461 |
| Q5 | 2.23 (1.75, 2.85) | < 0.001 |  | 1.52 (1.02, 2.29) | 0.042 |  | 1.03 (0.75, 1.40) | 0.869 |  | 2.04 (1.29, 3.26) | 0.003 |  | 1.90 (0.74, 5.29) | 0.198 |
| PRS-BMI |  |  |  |  |  |  |  |  |  |  |  |  |  |  |
| Q1 | reference |  |  | reference |  |  | reference |  |  | reference |  |  | reference |  |
| Q2 | 0.98 (0.81, 1.19) | 0.842 |  | 1.16 (0.84, 1.62) | 0.367 |  | 1.13 (0.88, 1.45) | 0.337 |  | 1.49 (1.06, 2.11) | 0.023 |  | 1.30 (0.58, 2.98) | 0.523 |
| Q3 | 1.11 (0.92, 1.35) | 0.254 |  | 1.01 (0.72, 1.41) | 0.950 |  | 0.94 (0.72, 1.21) | 0.617 |  | 1.19 (0.83, 1.71) | 0.342 |  | 1.01 (0.43, 2.38) | 0.981 |
| Q4 | 1.15 (0.95, 1.38) | 0.153 |  | 1.41 (1.02, 1.94) | 0.036 |  | 1.02 (0.79, 1.31) | 0.895 |  | 1.07 (0.74, 1.55) | 0.699 |  | 1.80 (0.84, 3.99) | 0.134 |
| Q5 | 1.14 (0.95, 1.37) | 0.169 |  | 1.19 (0.86, 1.65) | 0.298 |  | 1.07 (0.83, 1.38) | 0.598 |  | 1.38 (0.97, 1.95) | 0.072 |  | 1.06 (0.45, 2.50) | 0.889 |
| PRS-BF% |  |  |  |  |  |  |  |  |  |  |  |  |  |  |
| Q1 | reference |  |  | reference |  |  | reference |  |  | reference |  |  | reference |  |
| Q2 | 0.99 (0.82, 1.19) | 0.898 |  | 1.06 (0.78, 1.45) | 0.698 |  | 0.84 (0.65, 1.07) | 0.164 |  | 1.06 (0.77, 1.46) | 0.712 |  | 0.86 (0.38, 1.91) | 0.712 |
| Q3 | 0.89 (0.74, 1.08) | 0.232 |  | 0.84 (0.61, 1.17) | 0.302 |  | 0.81 (0.63, 1.04) | 0.099 |  | 0.78 (0.55, 1.10) | 0.162 |  | 0.85 (0.38, 1.89) | 0.696 |
| Q4 | 0.95 (0.79, 1.15) | 0.622 |  | 0.93 (0.68, 1.28) | 0.668 |  | 1.00 (0.78, 1.27) | 0.989 |  | 0.74 (0.52, 1.04) | 0.083 |  | 1.32 (0.64, 2.77) | 0.460 |
| Q5 | 1.11 (0.93, 1.33) | 0.246 |  | 0.92 (0.66, 1.26) | 0.599 |  | 0.89 (0.70, 1.14) | 0.353 |  | 0.94 (0.68, 1.30) | 0.712 |  | 0.79 (0.34, 1.80) | 0.583 |
| PRS-WHRadjBMI |  |  |  |  |  |  |  |  |  |  |  |  |  |  |
| Q1 | reference |  |  | reference |  |  | reference |  |  | reference |  |  | reference |  |
| Q2 | 1.03 (0.86, 1.23) | 0.772 |  | 1.00 (0.72, 1.40) | 0.995 |  | 0.76 (0.59, 0.98) | 0.034 |  | 1.22 (0.87, 1.71) | 0.262 |  | 1.67 (0.80, 3.62) | 0.181 |
| Q3 | 0.99 (0.82, 1.19) | 0.893 |  | 1.10 (0.79, 1.52) | 0.581 |  | 0.92 (0.72, 1.17) | 0.497 |  | 1.05 (0.74, 1.50) | 0.775 |  | 0.94 (0.40, 2.16) | 0.878 |
| Q4 | 0.98 (0.81, 1.18) | 0.803 |  | 1.16 (0.84, 1.60) | 0.385 |  | 0.91 (0.71, 1.16) | 0.451 |  | 1.17 (0.83, 1.65) | 0.377 |  | 0.72 (0.29, 1.75) | 0.478 |
| Q5 | 0.95 (0.79, 1.15) | 0.609 |  | 1.30 (0.95, 1.79) | 0.106 |  | 0.93 (0.73, 1.19) | 0.576 |  | 1.14 (0.81, 1.62) | 0.447 |  | 1.52 (0.70, 3.37) | 0.292 |

BF%, body fat percentage; BMI, body mass index; CHD, Coronary heart disease; CI, confidence interval; OR, odds ratio; PAD, Peripheral artery disease; PRS, polygenic risk score; WHRadjBMI, waist-to-hip ratio after adjusting for BMI

Adjusted for age, sex, smoking and drinking status.
